# Infant attachment does not depend on neonatal amygdala and hippocampal structure and connectivity

**DOI:** 10.1101/2023.12.07.23299615

**Authors:** Lorena Jiménez-Sánchez, Manuel Blesa Cabez, Kadi Vaher, Amy Corrigan, Michael J. Thrippleton, Mark E. Bastin, Alan J. Quigley, Sue Fletcher-Watson, James P. Boardman

## Abstract

Infant attachment is an antecedent of later socioemotional abilities, which can be adversely affected by preterm birth. The structural integrity of amygdalae and hippocampi are associated with attachment in childhood. We aimed to investigate associations between neonatal amygdalae and hippocampi structure and their whole-brain connections and attachment behaviours at nine months of age in a sample of infants enriched for preterm birth. In 133 neonates (median gestational age 32 weeks, range 22.14–42.14), we calculated measures of amygdala and hippocampal structure (volume, fractional anisotropy, mean diffusivity, neurite dispersion index, orientation dispersion index) and structural connectivity, and coded attachment behaviours (distress, fretfulness, attentiveness to caregiver) from responses to the Still-Face Paradigm at nine months. After multiple comparisons correction, there were no significant associations between neonatal amygdala or hippocampal structure and structural connectivity and attachment behaviours: standardised β values -0.23 to 0.21, adjusted p-values > 0.34. Findings indicate that the neural basis of infant attachment in term and preterm infants is not contingent on the structure or connectivity of the amygdalae and hippocampi in the neonatal period, which implies that it is more widely distributed in early life and or that network specialisation takes place in the months after hospital discharge.

**Highlights:**

- 133 infants had brain MRI and attachment data based on the Still-Face Paradigm.
- The structure of amygdalae and hippocampi and their brain networks was examined.
- Neonatal amygdalae/hippocampi structure did not associate with infant attachment.
- Infant attachment is not contingent on neonatal amygdala/hippocampal connectivity.

**Graphical abstract:** 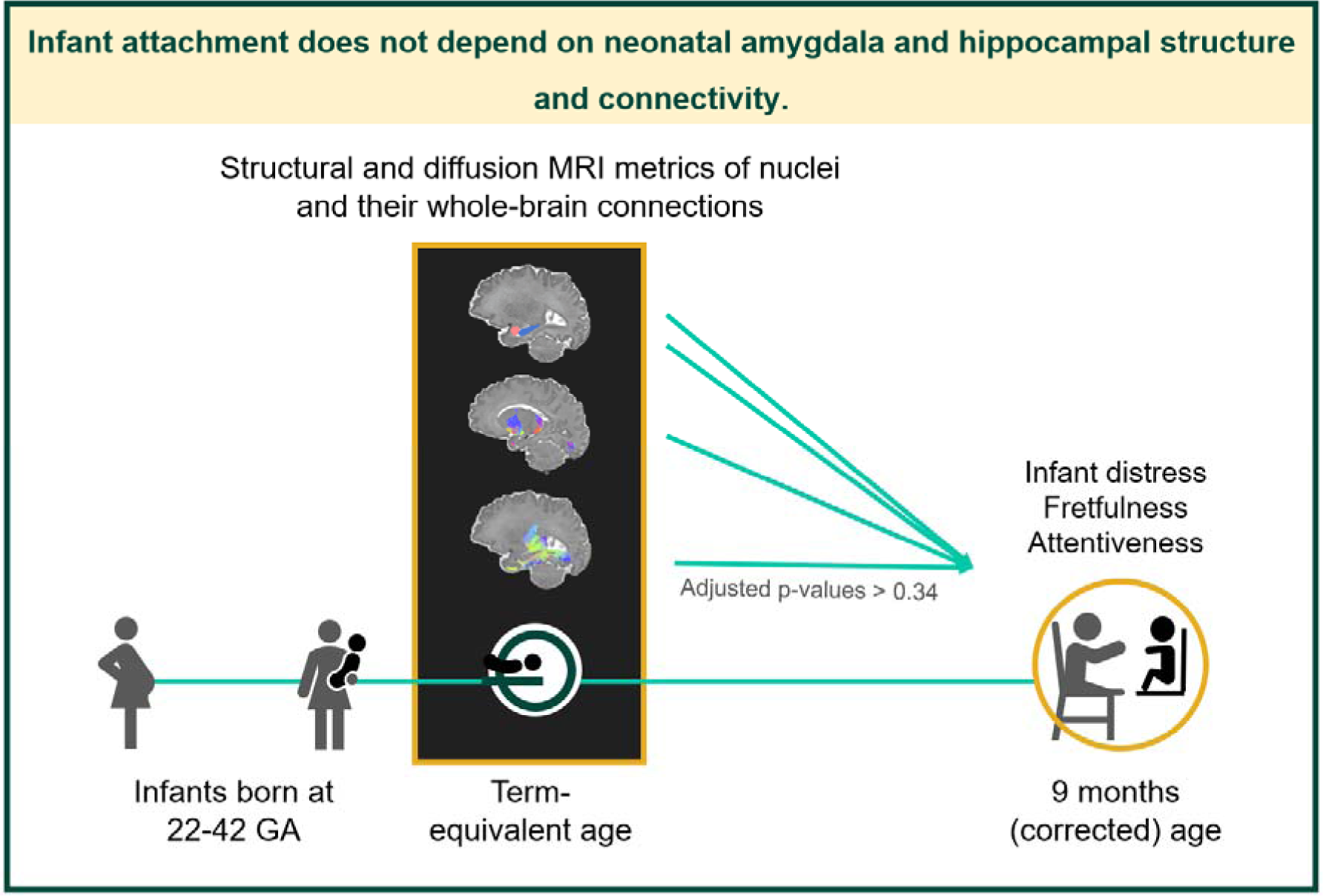

## 1. Introduction

In the first year of life, interactions between infants and their caregivers shape attachment behaviours (Ainsworth et al., 1972; Bowlby, 1982; Emde, 1980). Attachment can be conceived as a behavioural system that coordinates proximity and exploration to promote safety and independent learning. Infant behaviours within the attachment system (i.e. attachment behaviours) include proximity-seeking towards the caregiver and displaying distress, which helps draw the caregiver’s attention. An infant’s attachment style reflects qualitative differences in the manner in which the infant organises attachment behaviours towards a particular caregiver. Securely-attached infants use caregivers as a secure base to explore from and return to. Under a threat, infants with an insecure attachment style avoid their caregiver (avoidant), fail to respond independently (resistant) or exhibit contradictory behaviours i.e. attachment disorganisation (Ainsworth et al., 1981; Main & Solomon, 1990). Infant secure attachment positively associates with more effective emotion regulation (Cooke et al., 2019), and higher social competence (Groh et al., 2014) in later life. However, infant socioemotional development can also be impacted by adverse early life experiences. For example, preterm infants have higher likelihood to develop socioemotional difficulties, although there are uncertainties about whether low gestational age per se disrupts attachment (Arpi & Ferrari, 2013; Dean et al., 2021; Johnson & Marlow, 2011; Korja et al., 2012). Since attachment relationships precede socioemotional competence, identifying brain networks that underpin attachment and the factors that influence those networks may help inform interventions to foster socioemotional development.

Early contextual factors, such as aspects of parenting behaviour, only partly explain infant attachment (Madigan et al., 2006; Van Ijzendoorn et al., 1999), so it remains a question whether intrinsic infant characteristics support attachment behaviours that meet the goal of felt security. Individual differences in attachment behaviours may arise from how infants perceive and process parental support, threatening stimuli and regulate their emotions accordingly (Bernier & Meins, 2008). The limbic system develops earlier and faster than the cerebral cortex (Utsunomiya et al., 1999), and may prove especially important to process emotional experiences during infancy, while more diverse functional specialisation including cognitive control of emotion becomes established as children develop (Casey et al., 2008; Steinberg, 2017). Specifically, functional magnetic resonance imaging (MRI) studies suggest that the amygdala and hippocampus are important for fear processing in newborns (Graham et al., 2016; Thomas et al., 2019), and fear extinction (Fullana et al., 2018) and emotion regulation (Sergerie et al., 2008) in older populations. Moreover, the amygdala and hippocampal structure has been associated with the perinatal stress environment that could modulate early parent-infant interactions. For example, maternal hair cortisol concentration, a stable marker of chronic maternal hypothalamic-pituitary-adrenal axis activity in pregnancy, is associated with differences in the newborn’s amygdala microstructure and structural connectivity (Stoye et al., 2020). Differences in hippocampal structure influence the relationship between caregiving and attachment in early childhood: the interaction between neonatal left hippocampal volume and maternal sensitivity is associated with disorganised infant attachment (Rifkin-Graboi et al., 2019). Infant’s bilateral hippocampal volumes are associated with maternal sensitivity (Rifkin-Graboi et al., 2015), which predicts attachment security (De Wolff & Van Ijzendoorn, 1997). Importantly, the integrity of these nuclei and their networks may also be affected by preterm birth, as evidenced by the differences in hippocampal structure (Ball et al., 2012) and amygdala functional connectivity at rest (Rogers et al., 2017) apparent between term and preterm neonates at term-equivalent age.

Several studies have investigated longitudinal associations between parent-infant attachment relationships and brain structure in childhood (Bernier et al., 2019; Hidalgo et al., 2019; Leblanc et al., 2017), but the extent to which differences in brain structures around the time of birth associate with attachment within the first two years of life (Rifkin-Graboi et al., 2019; Tharner et al., 2011) is relatively understudied. This is important to understand for designing and implementing strategies to promote socioemotional development at the optimal time. These studies showed significant associations for the volume of the gangliothalamic ovoid, and the interaction between neonatal left hippocampal volume and maternal sensitivity (Rifkin-Graboi et al., 2019; Tharner et al., 2011), with attachment disorganisation. MRI volumetric data has been used to capture infants’ brain structural changes in relation to attachment, but volumetric changes reflect macrostructural physiological or pathological processes, including growth, injury and dysmaturation (Boardman & Counsell, 2020). Microstructural measures of brain nuclei and their connections enable further inference about neurite density and organisation, and may be useful for understanding how the development of brain networks supports attachment.

By combining neonatal multimodal brain MRI and infant behaviours coded from structured interactions with a caregiver, we aimed to investigate associations between neonatal amygdala and hippocampal structure or connectivity and attachment behaviours at nine months of age in a sample of infants enriched for preterm birth. We hypothesised that structural variation in these brain nuclei and their whole-brain connections at term-equivalent age associates with individual differences in attachment behaviours derived from infants’ responses to the Still-Face Paradigm at nine months of age: infants’ distress, fretfulness, and attentiveness to the caregiver.

## 2. Material and Methods

### 2.1 Participants

Participants were recruited to a prospective longitudinal cohort study, the Theirworld Edinburgh Birth Cohort (TEBC) (Boardman *et al*., 2020). Infants were born between September 2016 to September 2021 at the Royal Infirmary of Edinburgh, UK. Preterm infants transferred to the hospital *ex utero* for intensive care, and infants with major congenital malformation, chromosomal abnormality, congenital infection, cystic periventricular leukomalacia, haemorrhagic parenchymal infarction, and post-haemorrhagic ventricular dilatation were excluded. Informed written parental consent was obtained. Ethical approval was obtained from the National Research Ethics Service (16/SS/0154), South East Scotland Research Ethics Committee, and NHS Lothian Research and Development (2016/0255).

Collected information included infant sex (male/female), gestational age (GA) at birth, birthweight, self-reported maternal characteristics and the Scottish Index of Multiple Deprivation 2016 (SIMD) rank, generated from postcode information collected via parental questionnaire. SIMD rank (Scottish Government, 2016) is a multidimensional score generated by the Scottish government ranking localities’ deprivation according to local income, employment, health, education, geographic access to services, crime and housing.

### 2.2 MRI

#### Data Acquisition

MRI scans were performed at term-corrected gestation according to the study protocol (Boardman *et al*., 2020). Briefly, a Siemens MAGNETOM Prisma 3T MRI clinical scanner (Siemens Healthcare, Erlangen, Germany) and a 16- channel phased-array paediatric head coil were used to acquire: a 3D T2-weighted (T2w) sampling scheme with application-optimised contrasts using flip angle evolution structural scan (voxel size = 1mm isotropic), and axial diffusion MRI (dMRI) data. dMRI was acquired in two separate acquisitions to reduce the time needed to re- acquire any data lost to motion artefact. The first acquisition consisted of 8 baseline volumes (b = 0 s/mm2 [b0]) and 64 volumes (b = 750 s/mm^2^); the second consisted of 8 b0, 3 volumes with b = 200 s/mm^2^, 6 volumes with b = 500 s/mm^2^ and 64 volumes with b = 2500 s/mm^2^. In addition, an acquisition of 3 b0 volumes with an inverse phase encoding direction was performed. All dMRI images were acquired using single-shot spin-echo echo planar imaging with 2-fold simultaneous multislice and 2-fold in-plane parallel imaging acceleration and 2 mm isotropic voxels; all three diffusion acquisitions had the same parameters (TR/TE 3400/78.0 ms). Further details can be found elsewhere (Boardman *et al*., 2020).

Infants were fed, wrapped, and slept naturally. Flexible earplugs and neonatal earmuffs (MiniMuffs, Natus) were used for acoustic protection. Infants were monitored throughout, and scans were supervised by a doctor or nurse trained in neonatal resuscitation. Each acquisition was inspected contemporaneously for motion artefact and repeated if there had been movement while the baby was still sleeping; dMRI acquisitions were repeated if signal loss was seen in three or more volumes.

#### Data pre-processing

Structural images were reported by a radiologist with experience in neonatal MRI (AJQ). Visual inspection and quality control of raw structural and diffusion images were performed by experienced image analysts (MBC, KV). T2w images were processed using the developing Human Connectome Project (dHCP) minimal processing pipeline for neonatal data to obtain the bias field corrected T2w image, brain masks, tissue segmentation and label parcellation; tissue volumes were then calculated (Makropoulos et al., 2018).

dMRI processing was performed as previously described (Blesa et al., 2021; Stoye et al., 2020; Vaher et al., 2022). For each subject, the two dMRI acquisitions were concatenated and denoised using a Marchenko-Pastur-PCA-based algorithm (Veraart et al., 2016). The eddy current, head movement and EPI geometric distortions were corrected using outlier replacement and slice-to-volume registration (Andersson et al., 2017; Andersson et al., 2016; Andersson et al., 2003; Andersson & Sotiropoulos, 2016; Smith et al., 2004). Bias field inhomogeneity correction was performed by calculating the bias field of the mean B0 volume and applying the correction to all the volumes (Tustison et al., 2010). Neurite orientation dispersion and density imaging (NODDI) and diffusion tensor imaging (DTI) maps were calculated in the dMRI processed images to obtain fractional anisotropy (FA), mean diffusivity (MD), neurite density index (NDI), and orientation dispersion index (ODI). DTI model was fitted in each voxel using the weighted least-squares method DTIFIT as implemented in FSL using only the b = 750 s/mm^2^ shell. NODDI metrics of amygdalae and hippocampi were calculated using all shells and the recommended values for neonatal grey matter of the parallel intrinsic diffusivity (1.25 µm^2^/ms) using the original NODDI MATLAB toolbox. NODDI metrics of amygdalae and hippocampi whole-brain connections were calculated following the same approach, using the recommended values for neonatal white matter of the parallel intrinsic diffusivity (1.45 µm^2^/ms) (Guerrero et al., 2019; Zhang et al., 2012). Registration between dMRI and T2w was calculated using boundary-based registration (Greve & Fischl, 2009).

Nonlinear diffeomorphic multimodal registration with T2w and tissue probability maps using ANTs symmetric normalisation (SyN) (Avants et al., 2008) was then performed between the subject and the dHCP extended volumetric atlas (Fitzgibbon et al., 2020). The transformation was combined with the diffusion-to-structural transformation to obtain the final diffusion-to-template alignment.

#### Structural connectivity of amygdalae and hippocampi

Seed-based tractography from source regions of interest (ROIs, left and right hippocampi, and left and right amygdalae) to the whole brain was used to generate a whole-brain connectivity mask for each unilateral nucleus across subjects. The reason to take this approach in lieu of anatomically-constraint tractography was twofold. First, most relevant networks for attachment in human infancy are uncertain. Second, neonatal atlases do not currently include relevant regions with well-known connections to the hippocampus and amygdala, such as the hypothalamus (Nieuwenhuys et al., 2007), likely due to the resolution required to parcellate these areas in the neonatal brain. Seed-based tractography from source ROIs to the whole brain allowed a more unbiased inclusion of tracts between source ROIs and highly-connected areas of interest that may not be captured by current neonatal atlases, while reducing the number of multiple comparisons.

Seed-based tractography from source ROIs to the whole brain was performed using constrained spherical deconvolution (Smith et al., 2012; Tournier et al., 2019). The required 5-tissue type file, was generated by combining the tissue probability maps obtained from the dHCP pipeline with the subcortical structures derived from the parcellation process (Blesa et al., 2021). Multi-tissue response function was calculated, with a FA threshold of 0.10 (Dhollander et al., 2019; Dhollander et al., 2016). The average response functions were calculated. Then, the multi-tissue fibre orientation distribution (FOD) for white matter and cerebrospinal fluid were calculated (Jeurissen et al., 2014), and global intensity normalisation on the FODs images was performed. Finally, a tractogram was created, generating 500,000 streamlines from the seed ROI, setting the backtrack option while the rest of the parameters were used as by default.

To find a balance between filtering out noisy streamlines and keeping biologically-relevant connections from unilateral nuclei, *purifibre* (Aydogan) was applied to each participant to retain 50% of the fibres (Figure S1). Then these tracts were binarised, and the binarised tracts were propagated to the common template using the previously computed registration. Masks across subjects were averaged, and the final image was generated by keeping only voxels common to 50% of the subjects and propagating it back to each subject. This was the final mask of the whole-brain structural connectome used to calculate the mean values of the different metrics.

#### Summary of MRI metrics

In all, MRI metrics included volume, FA, MD, NDI and ODI of left and right amygdalae, and left and right hippocampi; FA, MD, NDI and ODI of whole-brain connections of left and right amygdalae, and left and right hippocampi.

### 2.3 Infant behavioural data

Caregivers were invited to attend follow-up appointments with their infants at nine months of age (corrected age for preterm infants) at a University of Edinburgh site at the Royal Edinburgh Hospital. The visit comprised an extensive assessment battery including questionnaires and behavioural procedures, as described in the study protocol (Boardman *et al*., 2020).

#### The Still-Face Paradigm

After consent for video-recording was obtained, infants were secured in a high chair. Caregivers seated facing the infant approximately 50 cm away. Two Panasonic HC-W580 video cameras were set up behind the caregiver and the infant to record the infant and caregiver’s faces and hands. The researcher was seated out of view, and verbally cued caregivers. The procedure included five two-minute episodes (Haley & Stansbury, 2003); modified from the original protocol (Tronick et al., 1978): baseline, still-face, reunion, still-face, and reunion. During the baseline episode, caregivers were instructed to interact naturally with the infant, without using toys. During still-face episodes, caregivers were instructed to express a neutral facial expression, remaining still and looking slightly above the infant’s head, avoiding eye contact and interaction with their infant. During reunion episodes, caregivers were instructed to interact normally with the infant again, without toys. Caregivers were always given the option to terminate the paradigm if infants exhibited severe distress.

#### Data coding

Williams and Turner’s coding scheme (Williams & Turner, 2020) was selected to study attachment from the SFP at nine months of age. Williams and Turner’s coding scheme includes the Happy-Distressed (HD), Not fretful-Fretful (NFF) and Attentive-Avoidant (AA) Global Rating Scales (Murray et al., 1996) to analyse infant behaviours during the reunion episode (Abbott, 2016; Williams & Turner, 2020). The coding scheme was applied as per the coding manual. To study attachment behaviours, the HD, NFF and AA raw scores were calculated per infant. These scores represented percentages of time infants engaged in HD, NFF or AA behaviours during the reunion episode; ranging from 0 (longer time displaying distress, fretful or avoidant behaviours according to the scale) to 1 (longer time displaying happy, not fretful or attentive behaviours to the caregiver according to the scale). Appendix S1 elaborates on the coding scheme, including the attachment style categorisation system that was not used in this work.

Importantly, scores derived from infant responses to the SFP mirror traditional attachment behaviours described by Bowlby (Bowlby, 1982) and Ainsworth (Abbott, 2016; Ainsworth et al., 2015) in the Strange Situation. In specific, avoidance of proximity and contact in the Strange Situation is comparable to the AA scale, although the latter only involves avoidance of eye contact (other avoidant behaviours cannot be considered since infants’ behavioural responses are constrained by sitting in a highchair in the SFP). Amount of crying in the Strange Situation is captured by the HD scale, which details the proportion of time spent in a distressed state during each episode. This is a dual ended scale and also includes markers of happiness or positive affect. Resistant behaviours in the Strange Situation are captured by the NFF scale, which considers fretful behaviours infants display when resisting or protesting such as angry vocalising, screaming, or arching the back away from the caregiver. Contact maintaining in the SFP is indirectly captured by the AA scale, along with positive affect. In other words, the infant is able to maintain contact with the caregiver by being attentive, engaged and positive towards the caregiver. Proximity seeking in the Strange Situation has no direct parallels in codes from the SFP since infants are unable to move towards or from the caregiver in the SFP (Abbott, 2016).

The SFP in the TEBC was designed to explore a broad range of infant behaviours, including infant stress responses which are more pronounced following repeated still-face episodes (Provenzi *et al*., 2016). Thus, a double-still face design was selected. In this study, only the first reunion episode of the five-episode SFP was coded because this provided a better equivalent to previous studies, where infants only experienced one stress episode (the still-face episode) before the reunion (Abbott, 2016; Williams & Turner, 2020). Participants were excluded based on SFP procedural violations initiated by others than the caregiver-infant dyad, and coded reunion time lower than 30 seconds (see Appendix S2 for further details). Videocoding was conducted using EUDICO Linguistic Annotator software (Wittenburg et al., 2006).

Reliability analyses were carried out for a larger sample of the study cohort (n = 167 infants, not all of whom contributed MRI data). Two types of reliability scores were calculated. For attachment dimensions (raw scores on the HD, NFF and AA scales), the intraclass correlation coefficient (ICC) was calculated. Following Williams and Turner’s coding scheme, two coders would need to agree on the absolute values of the HD, NFF, and AA raw scores to assign the same attachment style to a subject (Williams & Turner, 2020). Thus, a two-way random-effects agreement ICC was calculated for each scale. For attachment categories (attachment styles), the percentage of agreement (tolerance = 0) and Cohen’s Kappa were calculated. For inter-rater reliability analyses, the second coder coded a randomly-chosen 10% of all available videos, raw scores and attachment styles were extracted and compared. For intra-rater reliability analyses, the main coder coded a randomly-chosen 10% of all available videos again, raw scores and attachment styles were extracted and compared with the first time videos were coded.

### 2.4 Statistical analyses

Data analyses and plots were generated using R, version 4.3.2 (R Core Team, 2023). Relevant R packages are listed in Appendix S3.

To study associations between infant neuroanatomy and attachment behaviours, general linear regression was used with each scaled attachment behaviour (z-transformed HD, NFF, and AA raw scores) as outcome, each scaled adjusted brain structural measure as predictor, and covariates as additive predictors. To control for brain size and growth, derived MRI measures were adjusted for GA at scan prior to regression analyses by fitting a linear model of each z-transformed brain structural measure on GA at scan and retaining the residuals.

Possible confounding variables were explored from screened and/or included covariates in studies investigating bidirectional associations between neuroanatomy and observational measures of attachment in infancy or childhood (Hidalgo et al., 2019; Leblanc et al., 2017; Rifkin-Graboi et al., 2015; Tharner et al., 2011). To be controlled for, identified variables that may modify the relationship between infant neuroanatomy and attachment behaviours had to meet either the definition of a confounder or a confound blocker (Wysocki et al., 2022). As confounders, we considered SIMD rank, infant sex, infant GA at birth, maternal age, maternal education, maternal antenatal smoking, maternal antenatal alcohol consumption, maternal antenatal depression, and maternal antenatal anxiety (Appendix S4). Infant birthweight also met the definition of confounder, but we did not include it as a potential covariate because of high collinearity with GA at birth (rho_Spearman_ = 0.862). To investigate correlations between attachment behaviours or brain measures and continuous variables, we used Pearson or Spearman correlation coefficients as a nonparametric alternative. To compare attachment behaviours or brain measures between groups defined by categorical nominal variables (e.g., male and female infants), we used two-sample t-tests or Mann-Whitney-Wilcoxon test as a nonparametric alternative. Tests to identify covariates were exploratory, so reported p-values were not corrected for multiple comparisons (Armstrong, 2014). Variables that were significantly associated with at least one attachment behaviour (HD, NFF or AA raw score) and at least one brain measure at p-value < 0.05 were retained and controlled for. In analyses of amygdalae and hippocampi structure, we included maternal education and maternal antenatal anxiety as covariates (Table S1). In analyses of amygdalae and hippocampi structural connectivity, no covariates were included (Table S2). All reported *β* coefficients were standardised.

For regression analyses, the linearity of the association, homogeneity of variance, and normality of the model residuals were checked by inspecting the model diagnostic plots. Multicollinearity of predictor variables was calculated with the variance inflation factor (O’brien, 2007). Most regression models met the model assumptions. NFF raw score data was skewed, and regression models for the NFF raw scores showed issues with linearity of the association, normality of residuals, and homogeneity of variance. A logit transformation of NFF raw scores improved conformation to the assumptions of linearity of associations, and normality of model residuals as indicated by the diagnostic plots; homogeneity of variance was then confirmed by Goldfeld-Quandt tests. Since the direction of the results was similar when using transformed compared to untransformed NFF raw scores, results from analyses using untransformed data are reported in the main text. Results from analyses using NFF logit-transformed data are available in Table S3.

All reported p-values were adjusted for false-discovery rate using the Benjamini-Hochberg (BH) procedure within each family of experiments. Families of experiments were identified based on several assumptions. First, only the interaction of the left hippocampal volume and maternal sensitivity were found to be associated with infant attachment in a previous study (Rifkin-Graboi et al., 2015), so we expected lateralisation in the associations of amygdalae and hippocampi microstructure with infant attachment. Second, brain measures of nuclei structure mostly captured grey matter characteristics, whereas brain measures of whole-brain connections of nuclei captured white matter characteristics. Considering these different tissue configurations, grey matter and white matter metrics were expected to have different interpretations (Boardman & Counsell, 2020). Third, regression models for nuclei structure *versus* structural connectivity included different covariates. Fourth, all attachment behaviours (HD, NFF and AA raw scores) were derived from a single assessment (the SFP). Thus, eight families of experiments were identified: one for each unilateral nucleus’ structure, and for each unilateral nucleus’ whole-brain structural connectivity.

#### Post-hoc power analyses

The included sample provided 90% power to detect a small effect size of 0.11 in analyses of amygdalae and hippocampi structure, and 90 % power to detect a very small effect size of 0.08 in analyses of amygdalae and hippocampi structural connectivity (R packages are listed in Appendix S3).

## 3. Results

### 3.1 Baseline characteristics

One hundred and thirty-three infants had available neonatal MRI and SFP data (119 mother-infant dyads; 13 father-infant dyads, one grandmother-infant dyad, Figure S2). Median (range) GA at birth was 32^+0^ (22^+1^–42^+1^), birthweight was 2000 (370–4560) g, and SIMD rank was 4972 (137–6967). Demographic characteristics of the group are displayed in Table 1.

**Table 1.** Characteristics of the study sample.

|  | Caregiver-infant dyads<br>(n = 133) |
| --- | --- |
| Gestational age at birth / weeks, median (range) | 32<sup>+0</sup> (22<sup>+1</sup>–42<sup>+1</sup>) |
| Birthweight / grams, median (range) | 2000 (370–4560) |
| Preterm birth, Term:preterm | 64:69 |
| Gestational age at scan / weeks, mean (SD) | 41.06 (1.53) |
| Sex, M:F | 78:55 |
| Ethnicity (%)<sup>a</sup> |  |
| • White | 90.23 |
| • White and Asian | 3.01 |
| • Mixed | 3.01 |
| • White and Black African | 0.75 |
| • White and Black Caribbean | 0.75 |
| • Pakistani | 0.75 |
| Maternal age / years, mean (SD) | 33.20 (4.84) |
| Maternal education (%) |  |
| • None | 4.51 |
| • 1-4 National 5s / Standard Grades / GCSEs | 3.76 |
| • > 5 National 5s / Standard Grades / GCSEs | 2.26 |
| • A levels / Highers / equivalent | 18.05 |
| • College qualification (e.g. NC, HNC, HND) | 39.85 |
| • University undergraduate degree | 34.45 |
| • University postgraduate degree | 0.75 |
| Maternal smoking during pregnancy, none:any<sup>b</sup> | 129:4 |
| Maternal alcohol consumption during pregnancy, none:any<sup>c</sup> | 124:7 |
| Maternal antenatal depression, absence:presence | 113:20 |
| Maternal antenatal anxiety, absence:presence | 106:27 |
| SIMD rank, median (range) | 4513 (137–6967) |
| HD Raw scores, mean (SD) | 0.53 (0.21) |
| NFF Raw scores, median (range) | 1 (0.23–1) |
| AA Raw scores, mean (SD) | 0.31 (0.19) |

All maternal characteristics were self-reported measures. <sup>a</sup>Missing data: ethnicity data was not available for n = 2 participants, <sup>b</sup>missing data: maternal smoking during pregnancy data was not available for n = 3 participants, <sup>c</sup>missing data: maternal alcohol consumption during pregnancy data was not available for n = 2 participants. For missing data, percentages do not add up to 100 %. AA = Attentive-Avoidant scale; HD = Happy-Distressed scale; HNC = Higher National Certificate; HND = Higher National Diploma; GCSE: General Certificate of Secondary Education; NC = National Certificate, NFF = Not fretful-Fretful scale; SD = Standard Deviation; SIMD = Scottish Index of Multiple Deprivation 2016.

### 3.2 Segmentation and whole-brain connectivity masks of amygdalae and hippocampi

Amygdalae and hippocampi segmentations from a participant at term equivalent age viewed in native space are shown in **Error! Reference source not found.**A. The whole-brain connectivity mask of amygdalae mostly captured tracts between amygdala and the septal nuclei, lentiform nucleus, thalamus, hypothalamus, hippocampus, temporal and occipital lobe (Figure 1B). The whole-brain connectivity mask for the left hippocampus mostly captured tracts between hippocampus and the lentiform nucleus, hypothalamus, mesencephalon, insula and temporal lobe (Figure 1C).

**Figure 1.**
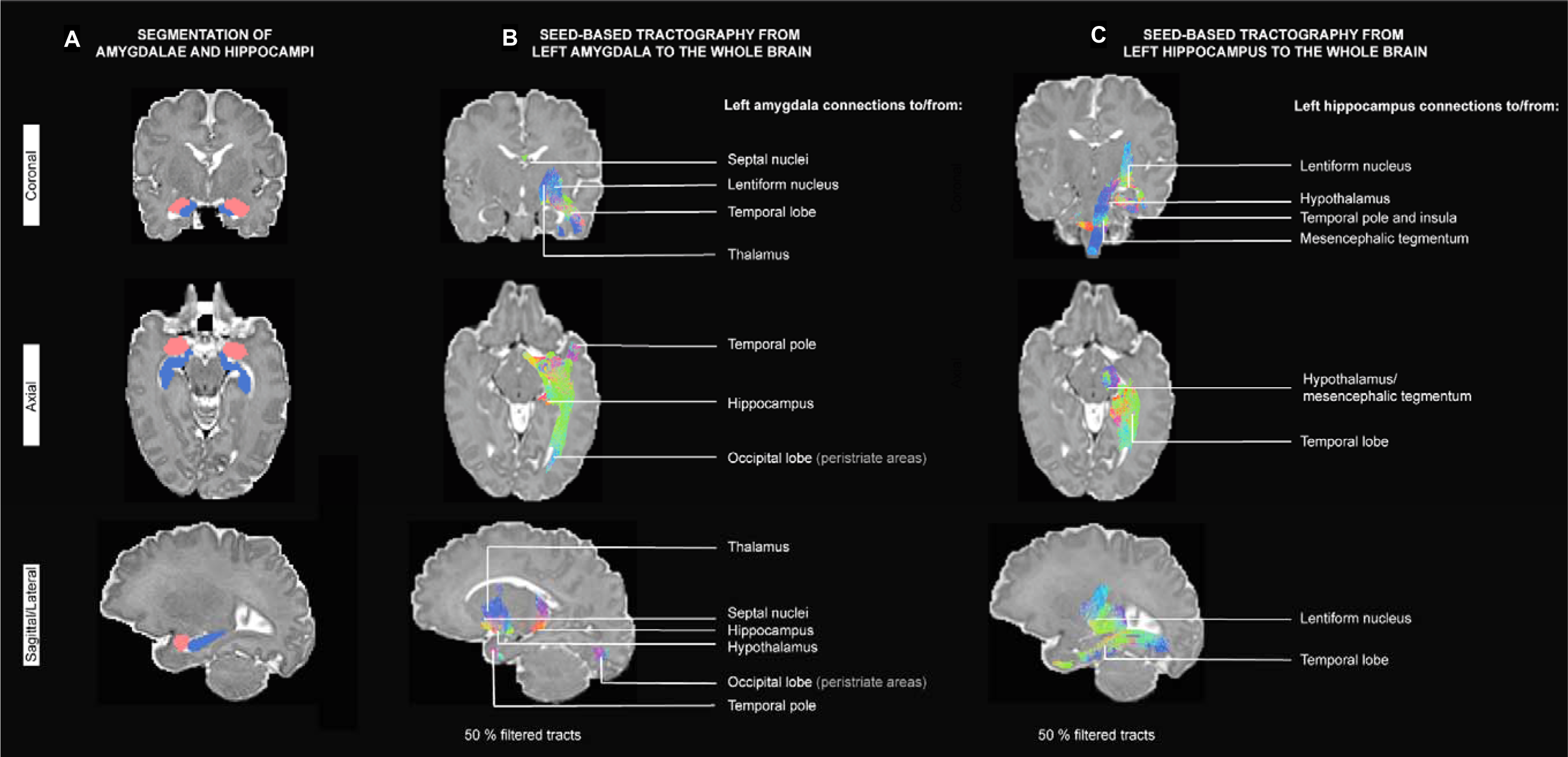
Segmentations and connections of interest in 50% filtered tracts from left amygdala and hippocampus (A) Amygdalae (red) and hippocampi (blue) segmentations from a participant at term-equivalent age, viewed in native space (left hemisphere is shown in sagittal/lateral sections). (B, C) Tractograms after filtering 50% of streamlines from a participant at term-equivalent age, viewed in native space. For visualisation purposes, afferent and efferent regions from each unilateral nucleus were inferred from previous anatomical descriptions of amygdalae and hippocampi structural connectivity (Nieuwenhuys et al., 2007).

### 3.3 Amygdalae and hippocampi macrostructure at birth and attachment behaviours at nine months of age

There were no significant associations between volumes of amygdalae and hippocampi and attachment behaviours (Figure 2A, Table 2).

**Figure 2.**
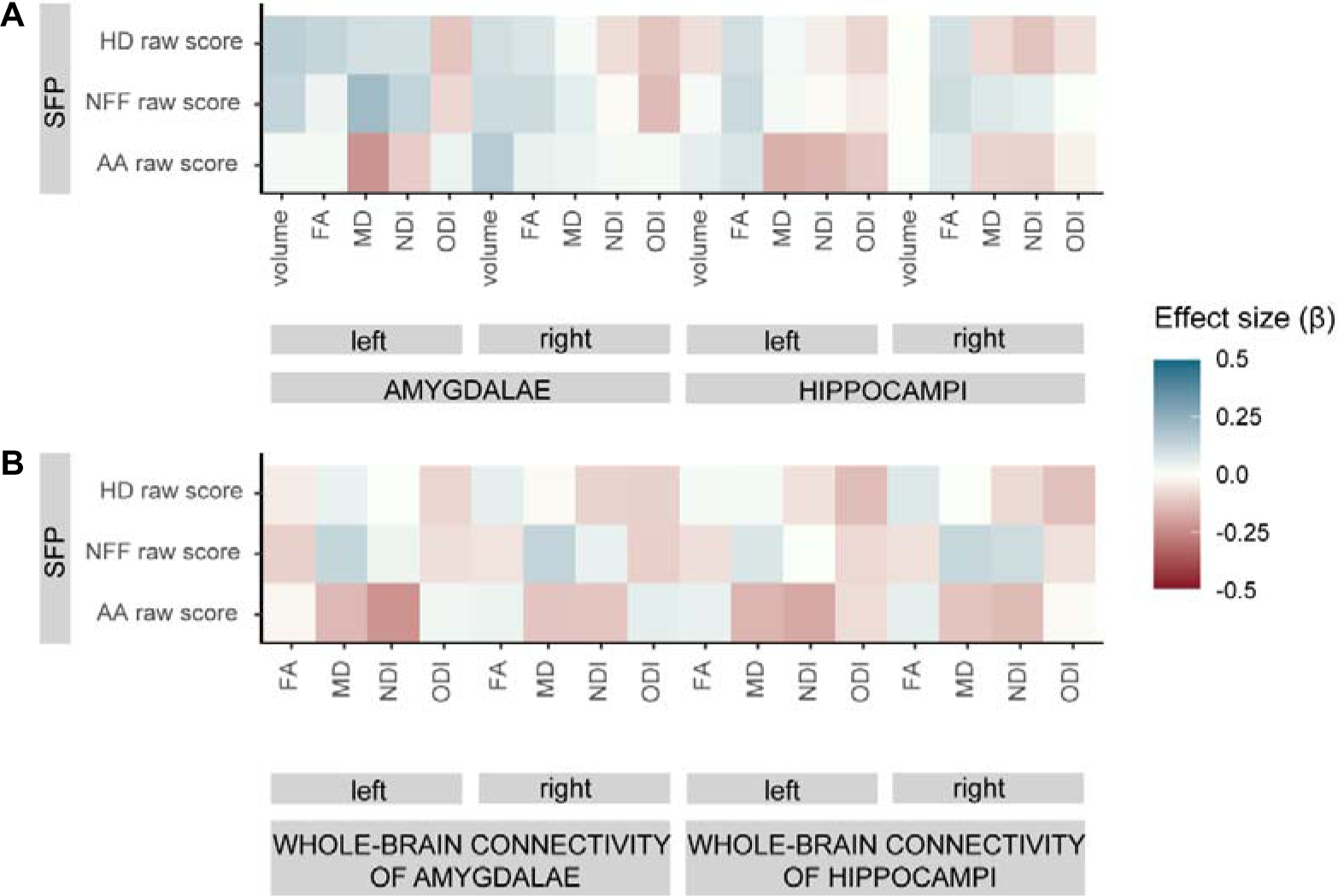
Associations between amygdalae and hippocampi structure and structural connectivity at birth and attachment behaviours in infancy. Colour indicates effect size (standardised β coefficient of regression models). Only corrected p-values are reported, see Table 2 for further details. AA = Attentive-Avoidant scale; FA = fractional anisotropy; HD = Happy-Distressed scale; MD = mean diffusivity; NDI = neurite density index; NFF = Not fretful-Fretful scale; ODI = orientation dispersion index.

**Table 2.**
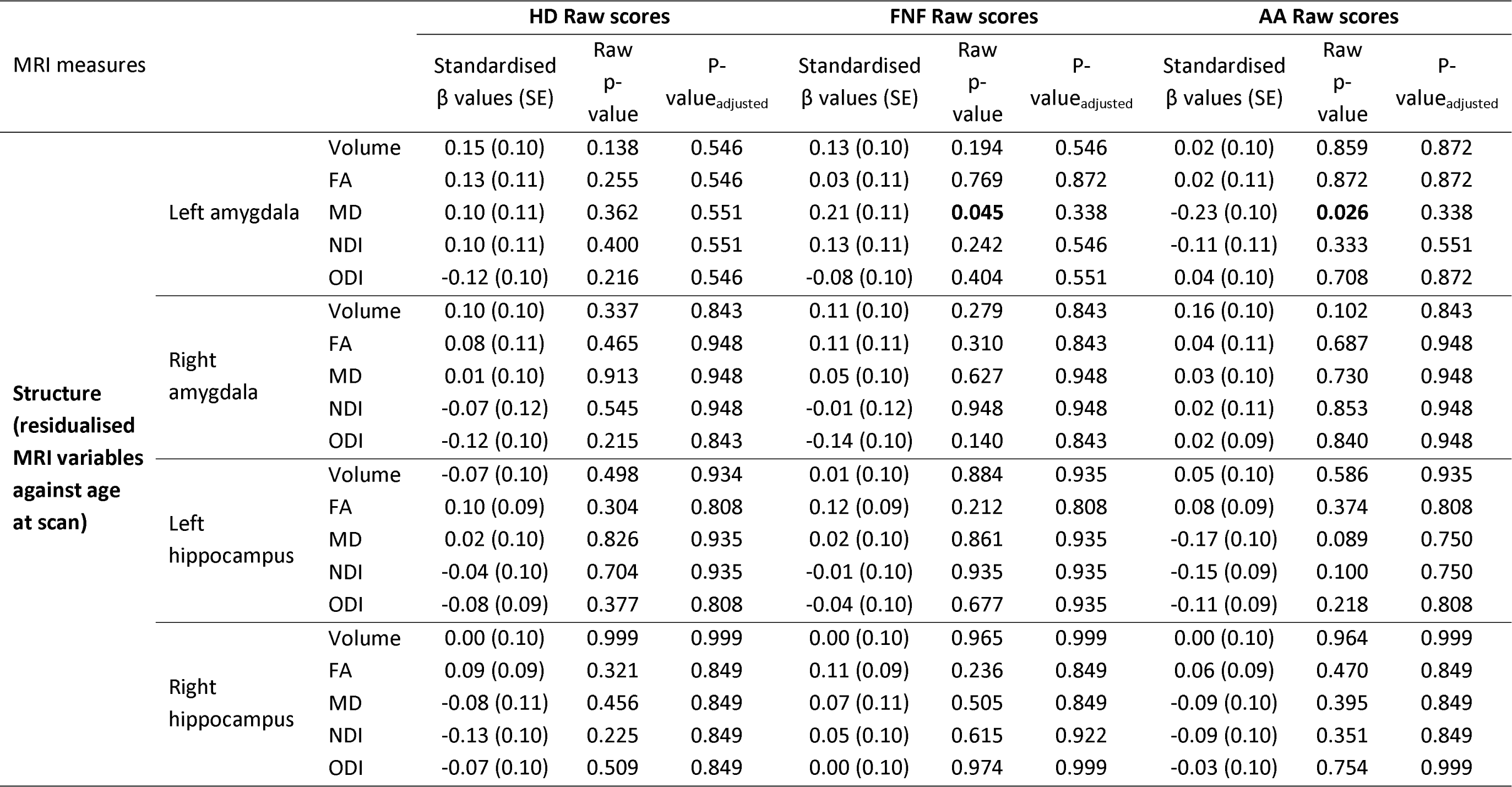
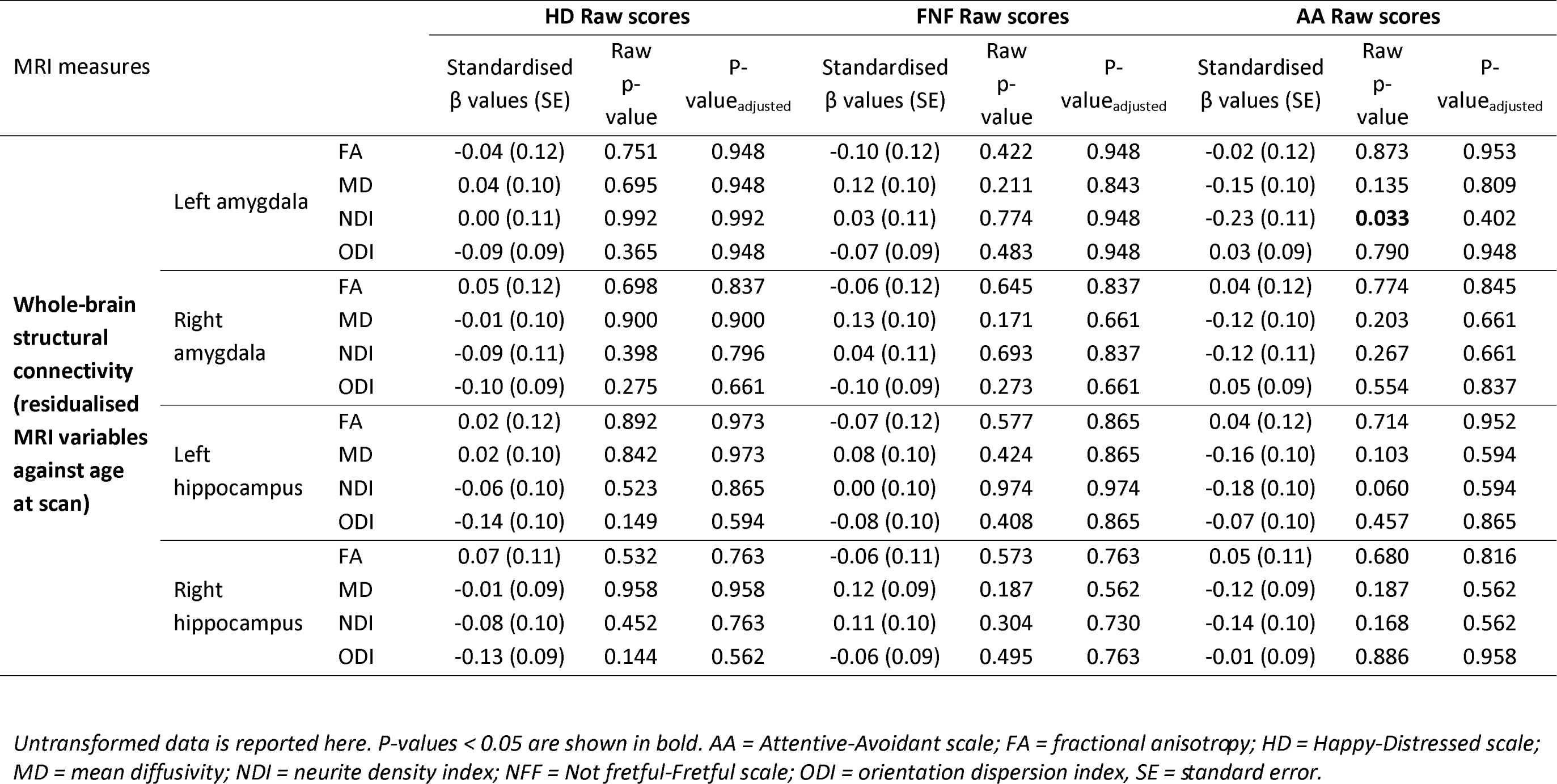
Associations between amygdalae and hippocampi structure and structural connectivity, and attachment behaviours.

### 3.4 Amygdalae and hippocampi microstructure at birth and attachment behaviours at nine months of age

There were nominally statistically significant associations between left amygdala MD and fretfulness, which did not survive correction for multiple comparisons (β (standard error, SE) = 0.21 (0.11), raw p-value = 0.045, p-value_adjusted_ = 0.338; Figure 2A, Table 2). This association was not nominally significant for transformed data (β (SE) = 0.20 (0.11), raw p-value = 0.059, Table S3).

There were nominally statistically significant associations between left amygdala MD and attentiveness to the caregiver, which did not survive correction for multiple comparisons (β (SE) = -0.23 (0.10), raw p-value = 0.026, p- value_adjusted_ = 0.338; Figure 2A, Table 2).

There were no other statistically significant associations between MRI metrics capturing microstructure of amygdalae or hippocampi and attachment behaviours (Figure 2A, Table 2).

### 3.5 Amygdalae and hippocampi structural connectivity at birth and attachment behaviours at nine months of age

There were nominally statistically significant associations between left amygdala whole-brain connectivity NDI and attentiveness to the caregiver, which did not survive correction for multiple comparisons (β (SE) = -0.23 (0.11), raw p-value = 0.033, p-value_adjusted_ = 0.402; Figure 2B, Table 2).

There were no other statistically significant associations between MRI metrics capturing the microstructure of amygdala or hippocampal whole-brain connections and attachment behaviours (Figure 2B, Table 2).

### 3.6 Reliability analyses for infant behavioural data

For inter-rater reliability analyses, ICC was 0.92 for the HD raw scores, 0.86 for the NFF raw scores and 0.81 for the AA raw scores. The percentage of agreement was 76.50% and Cohen’s kappa was 0.65 for attachment styles. For intra-rater reliability analyses, ICC was 0.82 for the HD raw scores, 0.85 for the NFF raw scores and 0.96 for the AA raw scores. The percentage of agreement was 82.40% and Cohen’s kappa was 0.69 for attachment styles.

## 4. Discussion

This study did not find statistically significant associations between the structure and structural connectivity of amygdalae and hippocampi around birth and attachment behaviours at nine months of age in a sample of infants born between 22 and 42 weeks of gestation and their primary caregivers. Therefore, the data do not support the hypothesis that structural variation in these brain nuclei and their whole-brain connections at term-equivalent age associates with individual differences in attachment behaviours derived from infants’ responses to the Still-Face Paradigm at nine months of age. It is possible that the structure of amygdalae and hippocampi is not a strong determinant of infant attachment in isolation, but may instead interact with environmental factors, such as maternal sensitivity, to predict attachment behaviour. This is supported by the reported significant interaction between neonatal left hippocampal volume and maternal sensitivity in predicting levels of infant disorganised behaviour, albeit in a smaller non-clinical sample (Rifkin-Graboi et al., 2019) compared with this study.

The results agree with the reported lack of associations between volumes of left and right amygdalae, or left and right hippocampi within the first two weeks of life and infant attachment at 1.5 years of age (Rifkin-Graboi et al., 2019). Here, we show that the finding applies to preterm infants, who are at risk of atypical brain development and socioemotional outcomes. Furthermore, we assessed microstructure and connectivity, in addition to macrostructure, evaluated attachment at a critical period of infant development, were powered to detect even small effects, and used a statistical approach that minimised the risk of error. To our knowledge, no study has explored associations between structural variation of amygdala or hippocampal networks and attachment in infancy or childhood. However, cross-sectional preliminary analyses showed maternal sensitivity positively associated with left hippocampal functional connectivity to regions important to social communication (left fusiform, left superior temporal cortex), and negatively associated with left hippocampal functional connectivity to regions important to memory (left entorhinal cortex) at six months of age (Rifkin-Graboi et al., 2015). This study raises the possibility that functional, rather than structural, characteristics of hippocampal networks play a role in the caregiver-infant relationship. A recent study indeed demonstrates that coupling between structural and functional connectivity is weaker in higher-level cognitive regions such as temporal areas (amygdala) compared to sensory regions (Preti & Van De Ville, 2019). Finally, other brain networks may more closely associate with infant attachment behaviours. For instance, the role of general cognitive ability in attachment has recently been highlighted (Del Giudice & Haltigan, 2023). It is thus possible that neural correlates of general cognitive ability, such as parieto-frontal networks (Basten et al., 2015; Hu et al., 2022; Langeslag et al., 2013), are more relevant for the development of attachment relationships than amygdala or hippocampal networks.

The structure of other brain nuclei at birth may contribute to attachment development later in infancy. For example, infants with a larger gangliothalamic ovoid measured using cranial ultrasound at six weeks had a lower risk of attachment disorganisation at 14 months, whereas volume of the lateral ventricles as an index of general brain development was not associated with attachment disorganisation in a large cohort of children with no neurological illness. However, the use of ultrasound limits inference about specific brain nuclei and networks that may support attachment (Tharner et al., 2011). While the basal ganglia are important for general goal-directed behaviour (Redgrave et al., 1999) and neurodevelopment (Boardman et al., 2010), the amygdala and hippocampus were candidate structures in this study because they more specifically support attachment processes due to their links with fear processing and extinction (Fullana et al., 2018; Graham et al., 2016; Thomas et al., 2019), and emotion regulation (Sergerie et al., 2008). Infant fear and emotion regulation play an essential role in attachment, as the presence of caregivers helps reduce fear and distress in the secure caregiver-infant relationship (Ainsworth et al., 1972; Hofer, 1994).

The strengths of this study include the use of a data-driven approach to study whole-brain connectivity that captured connections of the amygdala and hippocampus with structures in the midbrain, diencephalon and basal telencephalic structures; not frequently parcellated in current neonatal MRI atlases. We considered NODDI parameters to assess microstructure because ODI and NDI are functionally tractable (Kamiya et al., 2020). The potential role of the amygdala and hippocampus was assessed at term-equivalent age so possible impacts of caregiving after discharge from hospital on these structures were not expected. Based on prior literature, we explored a broad range of variables that may confound the association between neonatal brain measures and infant attachment and controlled for possible confounders. The relationships were described in a sample of infants enriched for preterm birth, and appear to apply across the whole GA range because GA at birth did not associate with both independent and dependent variables of this work (Tables S1 and S2). This suggests that the structural integrity of amygdalae and hippocampi and their whole-brain networks does not associate with later attachment behaviours in preterm and term infants. This work also has limitations. First, we did not use the Strange Situation (Ainsworth et al., 2015), the most commonly used assessment of infant attachment. Second, we did not consider caregiving behaviours such as maternal sensitivity, which predicts attachment security (De Wolff & Van Ijzendoorn, 1997) and could interact with brain neuroanatomy in predicting infant attachment (Rifkin-Graboi et al., 2019).

There are several research implications of this work. First, the structural neural substrates of attachment may include networks remote from the amygdala and hippocampi, and/or that these develop months after birth. These questions could be tested in future studies using longitudinal imaging throughout infancy. Second, if intrinsic infant factors around birth, such as neuroanatomy, play a negligible role in supporting attachment behaviours, interventions could focus on contextual factors of the infant postnatal environment, such as aspects of parenting behaviour, to cultivate infant attachment behaviours that positively associate with security and socioemotional development. Caregiving practices and infant social networks can vary significantly across cultures (Schmidt et al., 2021), so future studies could explore whether results of this work are generalisable to other populations, as well as other samples.

## 5. Conclusions

Structural variation in the amygdalae and hippocampi and their whole-brain connections at term-equivalent age do not contribute substantially to individual differences in attachment behaviours at nine months of age.

## Data Availability

Requests for anonymised data used in these analyses will be considered under the terms of the Theirworld Edinburgh Birth Cohort Data Access and Collaboration Policy: https://www.ed.ac.uk/centre-reproductive-health/tebc/about-tebc/for-researchers/data-access-collaboration.

## Acknowledgements

The authors are grateful to the families who participated in this research.

## 8. Data and code availability statement

Requests for anonymised data used in these analyses will be considered under the terms of the Theirworld Edinburgh Birth Cohort Data Access and Collaboration Policy: https://www.ed.ac.uk/centre-reproductive-health/tebc/about-tebc/for-researchers/data-access-collaboration. Scripts used for this analysis can be accessed at https://git.ecdf.ed.ac.uk/jbrl/amy-hip-attachment.

