## Supporting information for "Infant attachment does not depend on neonatal amygdala and hippocampal structure and connectivity"

### Appendix

**Appendix S1.** Williams and Turner’s coding scheme.

**Appendix S2.** Exclusion criteria.

**Appendix S3.** R packages used for statistical analyses.

**Appendix S4.** Potential covariates

**Table S1.** Amygdalae and hippocampi structure: covariate analysis.

**Table S2.** Amygdalae and hippocampi structural connectivity: covariate analysis.

**Table S3.** Associations between amygdalae and hippocampi structure and structural connectivity, and attachment behaviours: analyses including transformed data.

**Figure S1.** Seed-based tractography.

**Figure S2.** Study sample inclusion and exclusion flowchart.

**Appendix references.**

**Appendix S1**. Williams and Turner's coding scheme.

General coding guidelines:

1. Watch all phases of the Still-Face Paradigm in the video before coding
2. Only code the reunion phase, unless a disorganised attachment style is suspected.
3. Base all coding determinations only off of specific, observed behaviours listed in this manual.
4. If you are unsure, consult with another coder.

Coding procedure:

1. Counting second-by-second behaviours

The reunion phase of video should be watched three times. Each time, code a different dimension of behaviours. During seconds where the infant’s face cannot be seen, code the seconds as “unscorable”.

Happy-Distressed (HD) behaviours : infant’s codes for happy, neutral, or distressed are mutually exclusive. The infant must be assigned into one of three categories for every second of the reunion phase, unless the infant’s face is obscured from the camera. If unsure if infant is neutral or distressed, assign as “neutral.” If unsure is infant is happy or neutral, assign as “neutral”.

- “Happy” is defined by smiling, or positive vocalisations (laughing, giggling, cooing).
- “Neutral” is defined by a neutral expression or neutral vocalisations. Gaze on external stimuli, such as the mother, toys, chair, etc. with a neutral expression is counted as neutral.
- “Distressed” is defined by a negative expression on the face, negative vocalisations, whimpering, shouting, crying, fussing, kicking at caregiver, arching back in chair, batting at caregiver, pushing caregiver away, oral self-soothing (except a pacifier), hands clasping together, hiccups, spitting up, eyes glazing over and out of focus, or torso movements away from caregiver.

Not Fretful-Fretful (NFF) behaviours: for each second of the reunion phase, infant should be given a score of 1 in the NFF category if fretful or 0 if not fretful. Any behaviour that is not considered “fretful” is assigned “not fretful”. An infant can be distressed, but not fretful.

- “Fretful” is defined by arching the back, batting at the caregiver, pushing the caregiver away, “angry” vocalisations, shouting, kicking at caregiver, slamming hands down, or crinkling of face in “angry” affect.

Attentive-Avoidant (AA) behaviours: for each second of the reunion phase, infant gets a score of 1 in the AA category if attentive or 0 if avoidant. Any behaviour that is not considered “attentive” is assigned “avoidant”

- “Attentive” is defined by the infant looking in the direction of the caregiver’s face. Even if the caregiver is looking elsewhere, if the infant is looking at the caregiver’s face, the infant is scored as “attentive.”

2. Calculate percentages of time spent in behaviours and assign raw scores

HD: To calculate the HD raw score, count the number of total seconds of reunion time minus seconds of “unscorable” behaviour. This is your “coded reunion time.” Count the number of seconds in “happy”, “neutral”, and “distressed”. Divide the number of seconds in each category by “coded reunion time.” These are the percentages. Use the following equation to calculate the raw score: $HD raw score=\left( \% happy \right)+ \frac{(\% neutral)}{2}$

NFF: To calculate the NFF raw score, total the number of seconds scored “not fretful.” Divide them by the total number of seconds in “coded reunion time.”

AA: To calculate the AA raw score, total the number of seconds scored “attentive.” Divide them by the total number of seconds in “coded reunion time”.

3. Assign Global Rating Scales (GRS; 1-5) to HD, NFF and AA scores

For HD, NFF and AA raw scores, use the table below to assign each category a GRS score.

| GRS | Raw scores |
| --- | --- |
| 1 | < 0.200 |
| 2 | 0.200–0.399 |
| 3 | 0.400–0.600 |
| 4 | 0.601–0.800 |
| 5 | > 0.800 |

Note: For HD only, if infant receives a GRS score of “3”, a secondary GRS score is needed of one of the four options: 3(1), 3(2), 3(3), or 3(N). Use the following table for determining which score of “3” to assign: if infant meets criteria for 3(N) and another 3 score, final GRS score for HD would be 3(N).

| If scored a | |
| --- | --- |
| 66% or more of the distressed time in the first half of the reunion | 3(1) |
| 66% of the distressed time in the second half of the reunion | 3(2) |
| 41.1% -59.9% for both of the halves | 3(3) |
| if the infant is in the neutral category longer than 74% of the coded reunion time | 3(N) |

4. Match GRS scores to attachment algorithm and assign attachment status

| Classification | Reunion scales | | |
| --- | --- | --- | --- |
|  | Attentive-Avoidant | Happy-Distressed | Not fretful-Fretful |
| Disorganised | Low: 1–2 | Low: 1–2, 3(3)  3(2) on its own^a^ | Low: 1–3 |
| Resistant | Med/High: 3–5 |  | Low: 1–3 |
| Avoidant | Low: 1–2 | Med: 3(N), 3(1), 4 |  |
| Secure | Med/High: 3–5 | High: 3(N), 3(1), 3(3), 4–5 |  |

Note: Infant attachment style is shown on the left, and the three GRS scales (attentive-avoidant, happy-distressed, non-fretful-fretful) are on the right. aAn infant is automatically assigned a disorganised attachment style with a 3(2) score for Happy-Distressed, regardless of their scores on the AA and the Not fretful-fretful scales.

5. If score does not match algorithm, code still-face phase for HD, assign HD GRS score and re-assign attachment status

Occasionally, the three assigned GRS scores may not fit into the algorithm. In this case, the phase before the reunion phase, the still-face phase, needs to be considered before a final determination can be made. Count the number of seconds that the infant is “happy”, “neutral”, or “distressed” in the still-face phase, and derive the HD raw score the same way as done for the reunion phase, using the total seconds of scorable time from the still-face phase instead. If the infant has a higher raw HD score during the still-face phase than the reunion phase, assign a “disorganised” attachment style. Otherwise, infant receives an “unscorable” attachment style.

**Appendix S2**. Exclusion criteria.

We identified three main factors that could change the predictive behavioural responses of the infants during the Still Face Paradigm (SFP) or affect the video coding process:

- SFP procedural violations: During the SFP, caregivers or researchers could violate the procedure. Since only the first reunion episode was coded, only violations that happened before the first reunion episode were considered relevant for this study. During the first play or reunion episodes, violations included the use of objects not inherent to the face-to-face setting (pacifier, bottle or toys). During the still-face episode, violations included the caregiver turning away, touching or talking to the baby. Other relevant violations in this study included the infant’s early termination of the reunion episode (due to distress), interruptions by researchers or family members, researchers not concealed from view of the infant and caregiver, or infants being obscured by the camera.
- Wear of sensors: a subset sample of infants wore movement sensors on their wrists, ankles and torso as part of a sub-group analysis within the cohort protocol.
- Available coded reunion time: coded reunion time refers to the duration of the codable time during the reunion episode, in other words, the total number of seconds of the reunion time minus the number seconds of “unscorable” behaviour i.e. when the infant’s face was not visible (Williams & Turner, 2020). The available coded reunion time should be long enough to represent the infant’s response to the SFP.

Cases where infants could not terminate the reunion episode due to distress (early termination) or where the caregiver-infant dyad initiated the violation were considered to capture the clinical heterogeneity of the sample. Therefore, these participants were not excluded. However, cases where others than the caregiver-infant dyad i.e. researchers and other family members initiated the violation (researchers not concealed from view of the infant and caregiver, siblings present in room, etc.) were excluded. Mann-Whitney-Wilcoxon tests were used to test differences in attachment behaviours between infants wearing or not wearing sensors. Attachment behaviours were comparable in both groups (n = 24 infants of the final included sample wore sensors; p-values for HD, FNF and AA raw scores > 0.11), so no participants were excluded for this reason. Finally, the shortest duration of SFP episodes at nine months of age was 30 seconds in previous studies(Striano et al., 2005); a review on the duration of the SFP is published elsewhere(Mesman et al., 2009). Thus, cases where the coded reunion time was lower than 30 seconds were also excluded.

**Appendix S3.** R packages used for statistical analyses.

| Analysis | R package name | Citation |
| --- | --- | --- |
| Data manipulation | “dplyr” | (Wickham et al., 2019) |
|  | “stringr” | (Wickham & Wickham, 2019) |
|  | “tidytext” | (Silge & Robinson, 2016) |
|  | “tidyverse” | (Wickham & Wickham, 2017) |
| Plotting | “ggplot2” | (Wickham et al., 2016) |
|  | “ggpubr” | (Kassambara & Kassambara, 2020) |
|  | “scales” | (Wickham et al., 2022) |
| Power analysis | ‘WebPower’ | (Zhang et al., 2018) |
| Statistical analyses | ‘easystats’ | (Lüdecke et al., 2022) |
|  | “Hmisc” | (Harrell Jr & Harrell Jr, 2019) |
|  | “OpenMx” | (Boker, 2011 #587) |
|  | “performance” | (Lüdecke, Ben-Shachar, et al., 2021) |
|  | “rstatix” | (Kassambara, 2021) |
|  | “see” | (Lüdecke, Patil, et al., 2021) |
|  | “umx” | (Bates, 2019 #588) |
| Summary tables | “gtsummary” | (Daniel et al., 2021) |
|  | “vtable” | (Huntington-Klein, 2021) |
| Toolbox | “here” | (Müller, 2020 #589) |
|  | “htmltools” | (Cheng, 2023 #590) |
|  | “httpuv” | (Cheng, 2023 #593) |
|  | “knitr” | (Xie, 2023 #591) |
|  | “markdown” | (Xie, 2023 #592) |
|  | “NCmisc” | (Cooper, 2018) |
|  | “rlang” | (Henry & Wickham, 2020) |

**Appendix S4.** Potential covariates.

| Variable | Coded as | Type |
| --- | --- | --- |
| SIMD rank | 1 = most deprived to 6,976 = least deprived | Continuous |
| Infant sex | Male, female | Categorical nominal |
| Infant gestational age at birth | Weeks | Continuous |
| Maternal age | Years | Continuous |
| Maternal education (i.e. i.e. mother’s final educational qualification) | 1 = none, 2 = 1-4 National 5s / Standard Grades / General Certificate of Secondary Education, 3 = > 5 National 5s / Standard Grades / General Certificate of Secondary Education, 4 = A levels / Highers / equivalent, 5 = College qualification (e.g. National Certificate, Higher National Certificate, Higher National Diploma), 6 = University undergraduate degree, 7 = University postgraduate degree | Categorical ordinal |
| Maternal antenatal smoking | None, any | Categorical nominal |
| Maternal antenatal alcohol consumption | None, any | Categorical nominal |
| Maternal antenatal depression (self-reported) | Absence, presence | Categorical nominal |
| Maternal antenatal anxiety (self-reported) | Absence, presence | Categorical nominal (Wickham et al., 2016) |

**Table S1.** Amygdalae and hippocampi structure: covariate analysis.

| Continuous and categorical ordinal variables | | | **Gestational age at birth** | | **SIMD rank** | | **Maternal age** | | **Maternal education** | |
| --- | --- | --- | --- | --- | --- | --- | --- | --- | --- | --- |
|  |  |  | Correlation coefficient | p-  value | Correlation coefficient | p-  value | Correlation coefficient | p-  value | Correlation coefficient | p-  value |
| **Residualised MRI**  **variables against age at scan** | Left amygdala | Volume | 0.068 | 0.439 | 0.096 | 0.272 | 0.055^a^ | 0.532^a^ | 0.060 | 0.494 |
|  |  | FA | 0.163 | 0.062 | -0.064 | 0.463 | 0.010^a^ | 0.905^a^ | 0.042 | 0.629 |
|  |  | MD | 0.014 | 0.875 | -0.067 | 0.446 | -0.006^a^ | 0.946^a^ | 0.055 | 0.532 |
|  |  | NDI | 0.027 | 0.754 | -0.068 | 0.438 | 0.054^a^ | 0.538^a^ | 0.134 | 0.123 |
|  |  | ODI | 0.005 | 0.952 | 0.085 | 0.328 | 0.009^a^ | 0.917^a^ | 0.010 | 0.913 |
|  | Right amygdala | Volume | 0.018 | 0.838 | 0.002 | 0.978 | -0.003^a^ | 0.975^a^ | 0.105 | 0.227 |
|  |  | FA | 0.084 | 0.334 | 0.078 | 0.374 | -0.032^a^ | 0.715^a^ | -0.104 | 0.233 |
|  |  | MD | -0.088 | 0.317 | -0.052 | 0.550 | -0.034 | 0.696 | 0.063 | 0.472 |
|  |  | NDI | -0.080 | 0.358 | 0.057 | 0.512 | 0.019^a^ | 0.826^a^ | -0.048 | 0.584 |
|  |  | ODI | -0.036 | 0.682 | 0.000 | 0.998 | 0.100^a^ | 0.250^a^ | 0.059 | 0.498 |
|  | Left hippocampus | Volume | -0.068 | 0.436 | -0.013 | 0.883 | 0.022^a^ | 0.799^a^ | -0.023 | 0.792 |
|  |  | FA | 0.147 | 0.092 | 0.019 | 0.831 | -0.117^a^ | 0.180^a^ | 0.173 | **0.047** |
|  |  | MD | -0.206 | **0.017** | -0.073 | 0.406 | -0.052 | 0.555 | -0.011 | 0.901 |
|  |  | NDI | -0.272 | **0.002** | 0.028 | 0.751 | -0.128^a^ | 0.141^a^ | 0.053 | 0.542 |
|  |  | ODI | -0.202 | **0.019** | -0.010 | 0.911 | 0.095^a^ | 0.275^a^ | -0.075 | 0.389 |
|  | Right hippocampus | Volume | 0.012 | 0.890 | -0.072 | 0.408 | 0.091 | 0.297 | 0.077 | 0.378 |
|  |  | FA | 0.037 | 0.672 | -0.195 | **0.025** | -0.135^a^ | 0.122^a^ | -0.018 | 0.839 |
|  |  | MD | -0.401 | **0.000** | 0.043 | 0.627 | -0.089 | 0.307 | -0.096 | 0.270 |
|  |  | NDI | -0.349 | **0.000** | 0.044 | 0.616 | -0.052 | 0.548 | -0.017 | 0.849 |
|  |  | ODI | -0.081 | 0.354 | 0.012 | 0.889 | 0.057 | 0.513 | -0.047 | 0.588 |
| **SFP (behavioural) variables** | HD raw scores | | 0.116 | 0.185 | -0.033 | 0.706 | 0.034^a^ | 0.699^a^ | -0.012 | 0.888 |
|  | NFF raw scores | | -0.096 | 0.273 | -0.089 | 0.306 | -0.057 | 0.515 | -0.011 | 0.902 |
|  | AA raw scores | | 0.040 | 0.645 | 0.028 | 0.746 | 0.060^a^ | 0.491^a^ | -0.186 | **0.032** |

| Categorical nominal variables | | | **Sex** | | **Maternal smoking during pregnancy** | | **Maternal alcohol consumption during pregnancy**  **(none *versus* any)** | | **Maternal antenatal depression** | | **Maternal antenatal anxiety** | |
| --- | --- | --- | --- | --- | --- | --- | --- | --- | --- | --- | --- | --- |
|  |  |  | Statistic | p-value | Statistic | p-value | Statistic | p-value | Statistic | p-value | Statistic | p-value |
| **Residualised**  **MRI**  **Variables against age at scan** | Left amygdala | Volume | t = 3.51 | **0.001** | t = 1.06 | 0.357 | t = 0.13 | 0.901 | t = 2.94 | **0.006** | t = 0.71 | 0.482 |
|  |  | FA | t = -1.37 | 0.173 | t = -0.32 | 0.768 | t = 0.30 | 0.771 | t = -0.83 | 0.412 | t = -1.75 | 0.089 |
|  |  | MD | t = -0.43 | 0.667 | t = 0.16 | 0.884 | t = 0.27 | 0.798 | t = 0.36 | 0.722 | t = -0.33 | 0.747 |
|  |  | NDI | t = 0.04 | 0.971 | t = -0.02 | 0.986 | t = -0.03 | 0.980 | t = -0.38 | 0.705 | t = -1.05 | 0.301 |
|  |  | ODI | t = 0.14 | 0.887 | t = 1.27 | 0.282 | t = -1.18 | 0.277 | t = 1.07 | 0.292 | t = 2.20 | **0.034** |
|  | Right amygdala | Volume | t = 2.78 | **0.006** | t = 0.18 | 0.865 | t = 1.88 | 0.096 | t = 1.02 | 0.315 | t = 0.15 | 0.884 |
|  |  | FA | t = -1.05 | 0.295 | t = -0.23 | 0.835 | t = -0.7 | 0.507 | t = -0.74 | 0.466 | t = 0.22 | 0.826 |
|  |  | MD | W = 2320 | 0.425 | W = 285 | 0.661 | W = 423 | 0.914 | W = 1034 | 0.548 | W = 1259 | 0.336 |
|  |  | NDI | t = 2.33 | **0.021** | t = 1.50 | 0.171 | t = -0.86 | 0.417 | t = -0.24 | 0.810 | t = -0.67 | 0.503 |
|  |  | ODI | t = 0.79 | 0.433 | t = 0.20 | 0.850 | t = -0.40 | 0.701 | t = 1.02 | 0.316 | t = 2.32 | **0.025** |
|  | Left hippocampus | Volume | t = 2.01 | **0.046** | t = 0.43 | 0.692 | t = 0.26 | 0.802 | t = 0.91 | 0.373 | t = -0.51 | 0.614 |
|  |  | FA | t = 1.85 | 0.067 | t = 2.26 | 0.089 | t = 2.61 | **0.034** | t = 1.53 | 0.138 | t = -0.86 | 0.396 |
|  |  | MD | W = 2007 | 0.530 | W = 255 | 0.973 | W = 343 | 0.354 | W = 850 | 0.079 | W = 1539 | 0.548 |
|  |  | NDI | t = -0.09 | 0.927 | t = 0.01 | 0.992 | t = 0.54 | 0.606 | t = -1.55 | 0.134 | t = -0.33 | 0.744 |
|  |  | ODI | t = -1.16 | 0.247 | t = 0.90 | 0.432 | t = -3.05 | **0.018** | t = -0.45 | 0.656 | t = 0.64 | 0.527 |
|  | Right hippocampus | Volume | W = 2635 | **0.025** | W = 317 | 0.385 | W = 512 | 0.428 | W = 1052 | 0.626 | W = 1331 | 0.578 |
|  |  | FA | t = 1.38 | 0.169 | t = -0.63 | 0.569 | t = 0.17 | 0.872 | t = -1.87 | 0.072 | t = -1.39 | 0.171 |
|  |  | MD | W= 1886 | 0.238 | W = 198 | 0.471 | W = 193 | **0.014** | W = 840 | 0.068 | W = 1508 | 0.669 |
|  |  | NDI | W = 2393 | 0.258 | W = 207 | 0.549 | W = 379 | 0.577 | W = 876 | 0.111 | W = 1339 | 0.609 |
|  |  | ODI | W = 1984 | 0.463 | W = 297 | 0.549 | W = 359 | 0.446 | W = 979 | 0.343 | W = 1906 | **0.008** |
| **SFP**  **variables** | HD raw scores | | t = 0.41 | 0.683 | t = 2.67 | 0.063 | t = 0.18 | 0.863 | t = 1.20 | 0.239 | t = 0.97 | 0.338 |
|  | NFF raw scores | | W = 2242 | 0.622 | W = 393 | **0.033** | W = 435 | 0.995 | W = 1317 | 0.187 | W = 1501 | 0.662 |
|  | AA raw scores | | t = 1.05 | 0.297 | t = -0.84 | 0.460 | t = -0.54 | 0.607 | t = 0.62 | 0.540 | t = 2.38 | **0.022** |

*All correlations of continuous and categorical ordinal variables were performed using Spearman’s rho except otherwise specified. ^a^Correlations of maternal age performed using Pearson’s r. P-values < 0.05 are shown in bold. AA = Attentive-Avoidant scale; FA = fractional anisotropy; HD = Happy-Distressed scale; MD = mean diffusivity; NDI = neurite density index; NFF = Not fretful-Fretful scale; ODI = orientation dispersion index.*

**Table S2.** Amygdalae and hippocampi structural connectivity: covariate analysis.

| Continuous and categorical ordinal variables | | | **Gestational age at birth** | | **SIMD rank** | | **Maternal age** | | **Maternal education** | |
| --- | --- | --- | --- | --- | --- | --- | --- | --- | --- | --- |
|  |  |  | Correlation coefficient | p-  value | Correlation coefficient | p-value | Correlation coefficient | p-value | Correlation coefficient | p-  value |
| **Residualised MRI**  **variables against age at scan** | Left amygdala | FA | 0.219 | 0.011 | 0.057 | 0.518 | 0.001 | 0.988 | 0.115 | 0.187 |
|  |  | MD | -0.348 | **0.000** | -0.023 | 0.796 | -0.074 | 0.396 | 0.005 | 0.950 |
|  |  | NDI | -0.132 | 0.130 | 0.027 | 0.754 | -0.051^a^ | 0.559^a^ | 0.077 | 0.379 |
|  |  | ODI | -0.306 | **0.000** | 0.001 | 0.988 | 0.031 | 0.722 | -0.083 | 0.343 |
|  | Right amygdala | FA | 0.323 | **0.000** | 0.056 | 0.520 | 0.048 | 0.580 | 0.103 | 0.237 |
|  |  | MD | -0.385 | **0.000** | -0.015 | 0.864 | -0.083 | 0.340 | -0.006 | 0.944 |
|  |  | NDI | -0.109 | 0.210 | 0.090 | 0.303 | -0.049^a^ | 0.576^a^ | 0.027 | 0.754 |
|  |  | ODI | -0.254 | **0.003** | 0.005 | 0.957 | 0.044^a^ | 0.616^a^ | -0.097 | 0.265 |
|  | Left hippocampus | FA | 0.315 | **0.000** | -0.036 | 0.683 | 0.007 | 0.932 | 0.048 | 0.583 |
|  |  | MD | -0.247 | **0.004** | -0.027 | 0.761 | -0.009 | 0.918 | 0.018 | 0.837 |
|  |  | NDI | -0.100 | 0.250 | 0.025 | 0.774 | -0.012 | 0.891 | 0.057 | 0.514 |
|  |  | ODI | -0.323 | **0.000** | 0.058 | 0.508 | 0.051^a^ | 0.562^a^ | -0.041 | 0.639 |
|  | Right hippocampus | FA | 0.441 | **0.000** | -0.039 | 0.652 | 0.009 | 0.917 | 0.009 | 0.921 |
|  |  | MD | -0.327 | **0.000** | -0.032 | 0.712 | -0.046 | 0.600 | 0.010 | 0.909 |
|  |  | NDI | -0.146 | 0.095 | 0.086 | 0.327 | -0.062^a^ | 0.476^a^ | 0.024 | 0.785 |
|  |  | ODI | -0.340 | **0.000** | 0.086 | 0.325 | 0.090^a^ | 0.304^a^ | -0.031 | 0.725 |
| **SFP variables** | HD raw scores | | 0.116 | 0.185 | -0.033 | 0.706 | 0.034^a^ | 0.699^a^ | -0.012 | 0.888 |
|  | NFF raw scores | | -0.096 | 0.273 | -0.089 | 0.306 | -0.057 | 0.515 | -0.011 | 0.902 |
|  | AA raw scores  AA raw scores | | 0.040 | 0.645 | 0.028 | 0.746 | 0.060^a^ | 0.491^a^ | -0.186 | **0.032** |

| Categorical nominal variables | | | **Sex** | | **Maternal smoking during pregnancy** | | **Maternal alcohol consumption during pregnancy** | | **Maternal antenatal depression** | | **Maternal antenatal anxiety** | |
| --- | --- | --- | --- | --- | --- | --- | --- | --- | --- | --- | --- | --- |
|  |  |  | Statistic | p-value | Statistic | p-value | Statistic | p-value | Statistic | p-value | Statistic | p-value |
| **Residualised MRI variables against age at scan** | Left amygdala | FA | W = 2083 | 0.779 | W = 334 | 0.272 | W = 497 | 0.522 | W = 1295 | 0.300 | W = 1481 | 0.782 |
|  |  | MD | W = 2193 | 0.830 | W = 233 | 0.803 | W = 421 | 0.894 | W = 895 | 0.139 | W = 1529 | 0.587 |
|  |  | NDI | t = -2.27 | **0.026** | t = 1.24 | 0.295 | t = 0.64 | 0.545 | t = -0.92 | 0.369 | t = -1.03 | 0.311 |
|  |  | ODI | W = 2300 | 0.480 | W = 151 | 0.175 | W = 390 | 0.656 | W = 938 | 0.228 | W = 1531 | 0.578 |
|  | Right amygdala | FA | W = 1932 | 0.332 | W = 300 | 0.522 | W = 503 | 0.483 | W = 1358 | 0.152 | W = 1386 | 0.803 |
|  |  | MD | W = 2229 | 0.703 | W = 204 | 0.522 | W = 348 | 0.382 | W = 905 | 0.157 | W = 1501 | 0.697 |
|  |  | NDI | t = 1.99 | **0.049** | t = 1.47 | 0.208 | t = -0.27 | 0.795 | t = -0.39 | 0.700 | t = -1.22 | 0.229 |
|  |  | ODI | t = 1.56 | 0.121 | t = -0.60 | 0.588 | t = 0.41 | 0.695 | t = -0.48 | 0.635 | t = 0.35 | 0.732 |
|  | Left hippocampus | FA | W = 2247 | 0.643 | W = 327 | 0.315 | W = 499 | 0.509 | W = 1278 | 0.353 | W = 1475 | 0.808 |
|  |  | MD | W = 2108 | 0.868 | W = 196 | 0.454 | W = 384 | 0.612 | W = 855 | 0.084 | W = 1412 | 0.918 |
|  |  | NDI | W = 1772 | 0.089 | W = 344 | 0.217 | W = 420 | 0.890 | W = 1013 | 0.463 | W = 1337 | 0.601 |
|  |  | ODI | t = -0.86 | 0.390 | t = -0.89 | 0.432 | t = -0.04 | 0.972 | t = -0.43 | 0.673 | t = -0.59 | 0.556 |
|  | Right hippocampus | FA | W = 1983 | 0.461 | W = 304 | 0.487 | W = 530 | 0.328 | W = 1301 | 0.283 | W = 1365 | 0.714 |
|  |  | MD | W = 2165 | 0.931 | W = 199 | 0.475 | W = 350 | 0.393 | W = 931 | 0.211 | W = 1468 | 0.838 |
|  |  | NDI | t = -1.54 | 0.125 | t = 1.12 | 0.323 | t = 0.04 | 0.966 | t = -0.26 | 0.801 | t = 1.42 | 0.163 |
|  |  | ODI | t = 1.20 | 0.233 | t = -0.47 | 0.670 | t = 0.04 | 0.966 | t = 0.77 | 0.448 | t = 0.25 | 0.801 |
| **SFP**  **variables** | HD raw scores | | t = 0.41 | 0.683 | t = 2.67 | 0.063 | t = 0.18 | 0.863 | t = 1.20 | 0.239 | t = 0.97 | 0.338 |
|  | NFF raw scores | | W = 2242 | 0.622 | W = 393 | **0.033** | W = 435 | 0.995 | W = 1317 | 0.187 | W = 1501 | 0.662 |
|  | AA raw scores | | t = 1.05 | 0.297 | t = -0.84 | 0.460 | t = -0.54 | 0.607 | t = 0.62 | 0.540 | t = 2.38 | **0.022** |

*All correlations of continuous and categorical ordinal variables were performed using Spearman’s rho except otherwise specified. ^a^Correlations of maternal age performed using Pearson’s r. P-values < 0.05 are shown in bold. AA = Attentive-Avoidant scale; FA = fractional anisotropy; HD = Happy-Distressed scale; MD = mean diffusivity; NDI = neurite density index; NFF = Not fretful-Fretful scale; ODI = orientation dispersion index.*

**Table S3.** Associations between amygdalae and hippocampi structure and structural connectivity, and attachment behaviours including transformed data.

| MRI measures | | | **HD Raw scores** | | | **FNF Raw scores^a^** | | | **AA Raw scores** | | |
| --- | --- | --- | --- | --- | --- | --- | --- | --- | --- | --- | --- |
|  |  |  | Standardised β values (SE) | Raw p-value | P-value_adjusted_ | Standardised β values (SE) | Raw p-value | P-value_adjusted_ | Standardised β values (SE) | Raw p-value | P-value_adjusted_ |
| **Structure (residualised MRI variables against age at scan)** | Left amygdala | Volume | 0.15 (0.10) | 0.138 | 0.516 | 0.16 (0.10) | 0.109 | 0.516 | 0.02 (0.10) | 0.859 | 0.872 |
|  |  | FA | 0.13 (0.11) | 0.255 | 0.545 | 0.04 (0.11) | 0.738 | 0.852 | 0.02 (0.11) | 0.872 | 0.872 |
|  |  | MD | 0.10 (0.11) | 0.362 | 0.545 | 0.20 (0.11) | 0.059 | 0.445 | -0.23 (0.10) | **0.026** | 0.386 |
|  |  | NDI | 0.10 (0.11) | 0.400 | 0.545 | 0.15 (0.11) | 0.183 | 0.541 | -0.11 (0.11) | 0.333 | 0.545 |
|  |  | ODI | -0.12 (0.10) | 0.216 | 0.541 | -0.09 (0.10) | 0.380 | 0.545 | 0.04 (0.10) | 0.708 | 0.852 |
|  | Right amygdala | Volume | 0.10 (0.10) | 0.337 | 0.843 | 0.17 (0.10) | 0.098 | 0.512 | 0.16 (0.10) | 0.102 | 0.512 |
|  |  | FA | 0.08 (0.11) | 0.465 | 0.913 | 0.15 (0.11) | 0.177 | 0.644 | 0.04 (0.11) | 0.687 | 0.913 |
|  |  | MD | 0.01 (0.10) | 0.913 | 0.913 | 0.04 (0.10) | 0.733 | 0.913 | 0.03 (0.10) | 0.730 | 0.913 |
|  |  | NDI | -0.07 (0.12) | 0.545 | 0.913 | 0.02 (0.12) | 0.867 | 0.913 | 0.02 (0.11) | 0.853 | 0.913 |
|  |  | ODI | -0.12 (0.10) | 0.215 | 0.644 | -0.17 (0.10) | 0.084 | 0.512 | 0.02 (0.09) | 0.840 | 0.913 |
|  | Left hippocampus | Volume | -0.07 (0.10) | 0.498 | 0.755 | 0.07 (0.10) | 0.508 | 0.755 | 0.05 (0.10) | 0.586 | 0.755 |
|  |  | FA | 0.10 (0.09) | 0.304 | 0.755 | 0.15 (0.09) | 0.107 | 0.537 | 0.08 (0.09) | 0.374 | 0.755 |
|  |  | MD | 0.02 (0.10) | 0.826 | 0.826 | -0.04 (0.10) | 0.683 | 0.755 | -0.17 (0.10) | 0.089 | 0.537 |
|  |  | NDI | -0.04 (0.10) | 0.704 | 0.755 | -0.05 (0.10) | 0.616 | 0.755 | -0.15 (0.09) | 0.100 | 0.537 |
|  |  | ODI | -0.08 (0.09) | 0.377 | 0.755 | -0.06 (0.10) | 0.524 | 0.755 | -0.11 (0.09) | 0.218 | 0.755 |
|  | Right hippocampus | Volume | 0.00 (0.10) | 0.999 | 0.999 | 0.04 (0.10) | 0.702 | 0.870 | 0.00 (0.10) | 0.964 | 0.999 |
|  |  | FA | 0.09 (0.09) | 0.321 | 0.870 | 0.11 (0.09) | 0.248 | 0.870 | 0.06 (0.09) | 0.470 | 0.870 |
|  |  | MD | -0.08 (0.11) | 0.456 | 0.870 | 0.06 (0.11) | 0.593 | 0.870 | -0.09 (0.10) | 0.395 | 0.870 |
|  |  | NDI | -0.13 (0.10) | 0.225 | 0.870 | 0.04 (0.10) | 0.695 | 0.870 | -0.09 (0.10) | 0.351 | 0.870 |
|  |  | ODI | -0.07 (0.10) | 0.509 | 0.870 | -0.04 (0.10) | 0.725 | 0.870 | -0.03 (0.10) | 0.754 | 0.870 |
| **Whole-brain structural connectivity (residualised MRI variables against age at scan)** | Left amygdala | FA | -0.04 (0.12) | 0.751 | 0.948 | -0.06 (0.12) | 0.644 | 0.948 | -0.02 (0.12) | 0.873 | 0.953 |
|  |  | MD | 0.04 (0.10) | 0.695 | 0.948 | 0.11 (0.10) | 0.284 | 0.843 | -0.15 (0.10) | 0.135 | 0.809 |
|  |  | NDI | 0. (0.11) | 0.992 | 0.992 | 0.02 (0.11) | 0.857 | 0.948 | -0.23 (0.11) | **0.033** | 0.402 |
|  |  | ODI | -0.09 (0.09) | 0.365 | 0.948 | -0.07 (0.09) | 0.440 | 0.948 | 0.03 (0.09) | 0.790 | 0.948 |
|  | Right amygdala | FA | 0.05 (0.12) | 0.698 | 0.837 | -0.06 (0.12) | 0.611 | 0.837 | 0.04 (0.12) | 0.774 | 0.845 |
|  |  | MD | -0.01 (0.10) | 0.900 | 0.900 | 0.15 (0.10) | 0.121 | 0.661 | -0.12 (0.10) | 0.203 | 0.661 |
|  |  | NDI | -0.09 (0.11) | 0.398 | 0.796 | 0.04 (0.11) | 0.728 | 0.837 | -0.12 (0.11) | 0.267 | 0.661 |
|  |  | ODI | -0.1 (0.09) | 0.275 | 0.661 | -0.09 (0.09) | 0.337 | 0.661 | 0.05 (0.09) | 0.554 | 0.837 |
|  | Left hippocampus | FA | 0.02 (0.12) | 0.892 | 0.973 | -0.04 (0.12) | 0.745 | 0.865 | 0.04 (0.12) | 0.714 | 0.952 |
|  |  | MD | 0.02 (0.10) | 0.842 | 0.973 | 0.05 (0.10) | 0.574 | 0.865 | -0.16 (0.10) | 0.103 | 0.594 |
|  |  | NDI | -0.06 (0.10) | 0.523 | 0.865 | -0.03 (0.10) | 0.794 | 0.974 | -0.18 (0.10) | 0.060 | 0.594 |
|  |  | ODI | -0.14 (0.10) | 0.149 | 0.594 | -0.07 (0.10) | 0.454 | 0.865 | -0.07 (0.10) | 0.457 | 0.865 |
|  | Right hippocampus | FA | 0.07 (0.11) | 0.532 | 0.763 | -0.05 (0.11) | 0.665 | 0.763 | 0.05 (0.11) | 0.680 | 0.816 |
|  |  | MD | -0.01 (0.09) | 0.958 | 0.958 | 0.14 (0.09) | 0.145 | 0.562 | -0.12 (0.09) | 0.187 | 0.562 |
|  |  | NDI | -0.08 (0.10) | 0.452 | 0.763 | 0.09 (0.10) | 0.386 | 0.730 | -0.14 (0.10) | 0.168 | 0.562 |
|  |  | ODI | -0.13 (0.09) | 0.144 | 0.562 | -0.05 (0.09) | 0.595 | 0.763 | -0.01 (0.09) | 0.886 | 0.958 |

*^a^FNF raw scores were logit transformed. P-values < 0.05 are shown in bold. AA = Attentive-Avoidant scale; BH = Benjamini-Hochberg; FA = fractional anisotropy; HD = Happy-Distressed scale; MD = mean diffusivity; NDI = neurite density index; NFF = Not fretful-Fretful scale; ODI = orientation dispersion index; SE = standard error.*

**Figure S1.** Seed-based tractography.

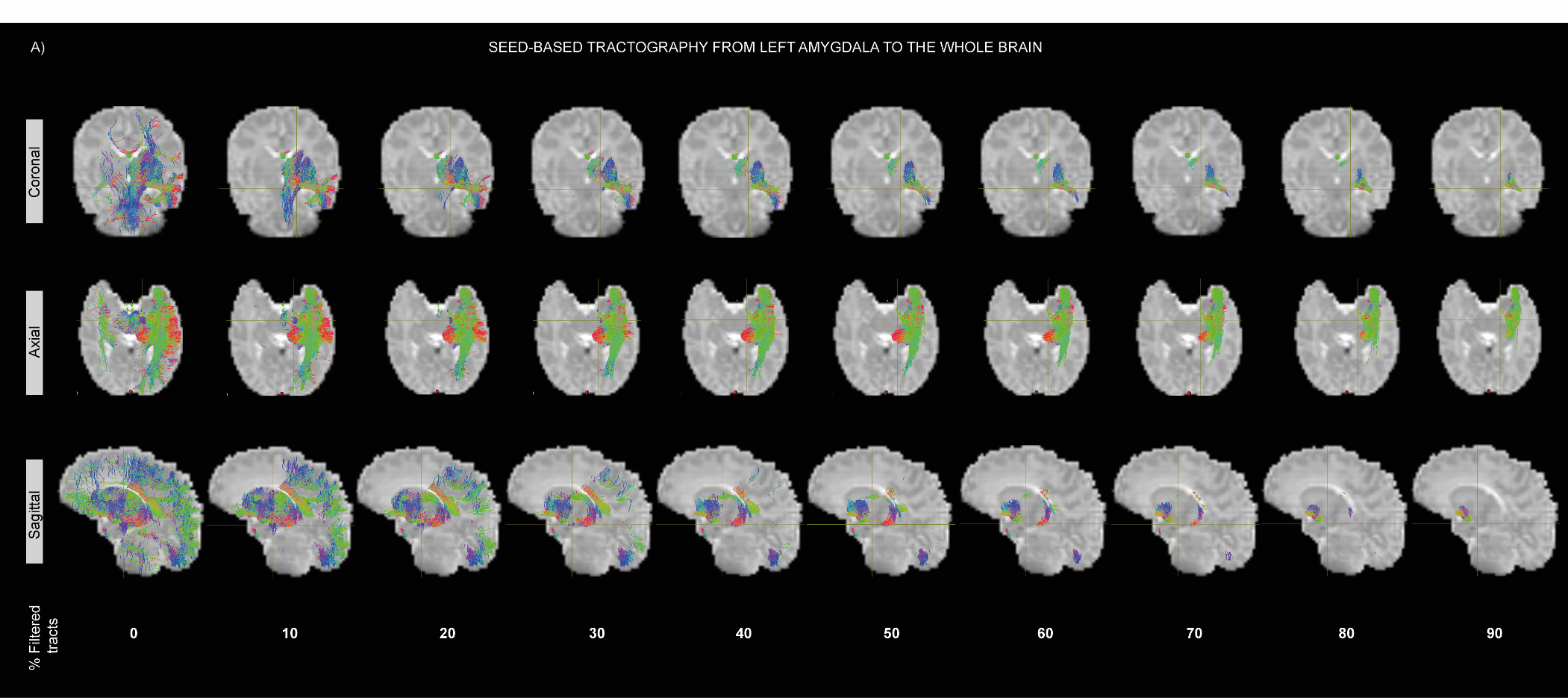

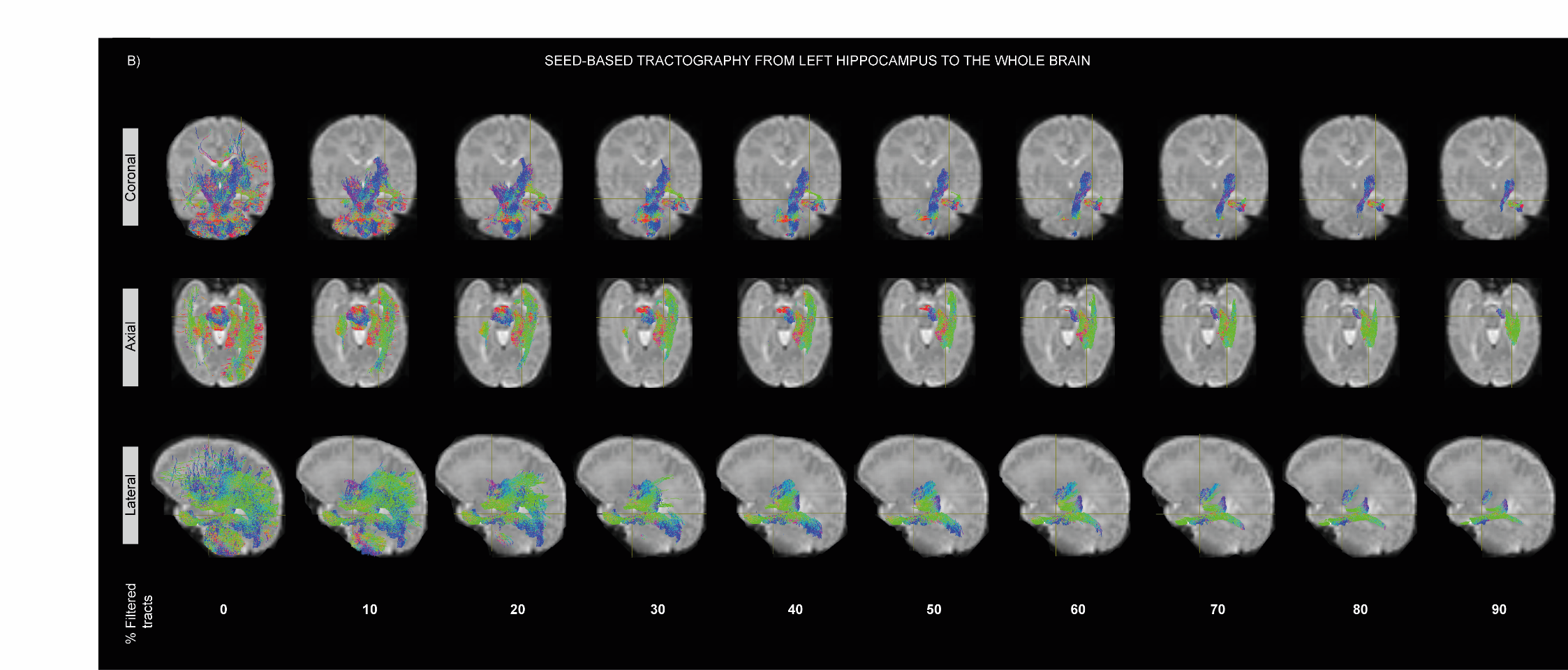

*Original and tractograms after filtering different percentages of streamlines are shown on the B0 from one participant (left hemisphere is shown in sagittal/lateral sections). Brown reference lines on each individual image indicate the location of coronal, axial and sagittal/lateral sections.*

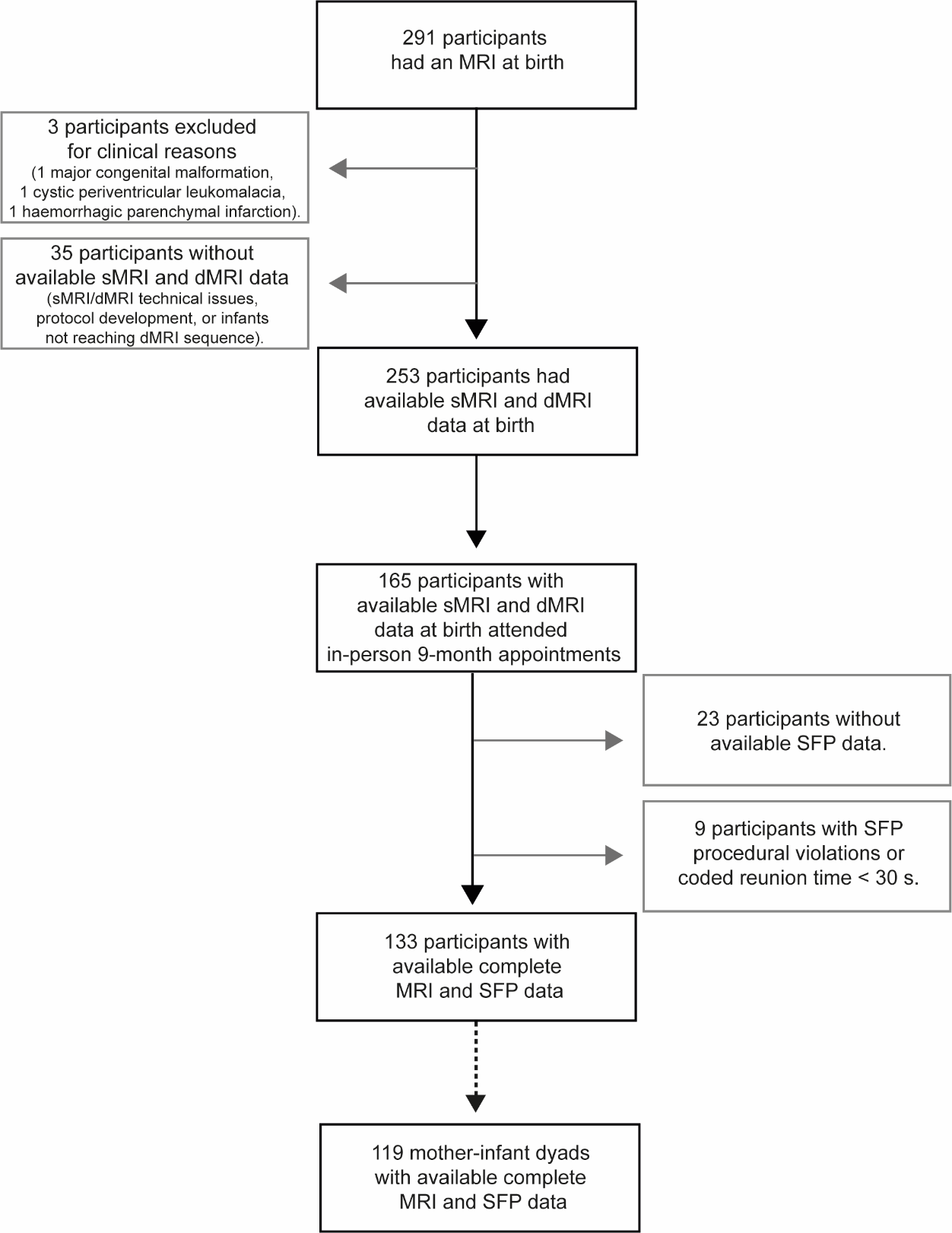

**Figure S2.** Study sample inclusion and exclusion flowchart.

*dMRI = diffusion magnetic resonance imaging; MRI = magnetic resonance imaging; SFP = Still-Face Paradigm; sMRI = structural magnetic resonance imaging.*
